# Hypertension in People Living with HIV (PLHIV): A Comparative Analysis Before and After Test-and-Treat Policy Implementation

**DOI:** 10.1101/2025.06.28.25330479

**Authors:** Martin Chakulya, David Chisompola, Siame Lukundo, Joreen P. Povia, Benson M. Hamooya, Annet Kirabo, Sepiso K. Masenga

## Abstract

**Background:** The universal test-and-treat (T&T) policy has improved HIV outcomes but may influence hypertension (HTN) risk due to prolonged antiretroviral therapy (ART) exposure. We compared HTN prevalence and risk factors among PLHIV before and after T&T implementation in Zambia.

**Methods:** A retrospective cohort study analyzed 6,409 PLHIV (2,920 pre-T&T and 3,489 post-T&T) from 12 Southern Province districts. Data on demographics, ART regimens, blood pressure, and laboratory measures were extracted from electronic (SmartCare) and paper records. Multivariable logistic regression identified HTN-associated factors (p<0.05).

**Results:** HTN prevalence increased from 8.8% pre-T&T to 10.2% post-T&T. Each year of age increased HTN odds by 5–6% in both cohorts (adjusted odds ratio [AOR]: 1.06 pre-T&T, 1.05 post-T&T p<0.0001). Urban residence was protective (AOR: 0.72 pre-T&T, 0.67 post-T&T p≤0.041), while males had higher HTN risk than females (12.2% vs. 8.8% post-T&T p=0.0015). Post-T&T, higher hemoglobin marginally increased HTN odds (AOR: 1.08; p=0.049). INSTI-based regimens rose from 26.3% to 41.5% post-T&T but showed no significant association with hypertension on multivariate analysis. Rural residents had higher HTN prevalence (11.5% vs. 8.4% urban post-T&T p=0.0027).

**Conclusions:** HTN prevalence increased post-T&T, and was driven by aging and potentially ART-related metabolic effects. Urban residence was unexpectedly protective, possibly due to better healthcare access. The hemoglobin-HTN link post-ART warrants further study. Strengths include a large, representative sample, though unmeasured confounders (e.g., lifestyle factors) and retrospective design limit causal inferences. Integrated HTN screening in HIV programs is critical to address this growing burden.

## Introduction

The widespread adoption of universal Test-and-Treat (T&T) policies for HIV has transformed the clinical landscape of antiretroviral therapy (ART), significantly reducing AIDS-related mortality and transmission [1]. However, as people living with HIV (PLWH) experience near-normal life expectancy, a new challenge has emerged the rising burden of non-communicable diseases (NCDs), particularly hypertension (HTN) [2,3]. While the benefits of early ART initiation such as immune preservation and viral suppression are well established, the long-term cardiovascular consequences of prolonged HIV infection and ART exposure remain understudied, especially in sub-Saharan Africa (SSA), where health systems remain disproportionately strained [3–5].

Before the T&T era, HTN in PLWH was often overlooked, as clinical priorities centered on managing opportunistic infections and delaying ART until CD4 counts declined [6]. Yet, even then, chronic immune activation and inflammation from untreated HIV likely contributed to endothelial dysfunction and accelerated atherosclerosis [7]. With the shift to immediate ART under T&T policies (post-2016), PLWH now face extended exposure to both ART-related metabolic effects (e.g., dyslipidemia, insulin resistance) and traditional HTN risk factors (e.g., aging, obesity, poor diet) [8]. Early evidence suggests that HTN prevalence among PLWH in SSA now exceeds that of the general population, yet screening and management remain inconsistent in HIV care programs [9,10].

The evolving epidemiology of hypertension (HTN) among people living with HIV (PLWH) before and after the implementation of Test-and-Treat (T&T) policies raises critical questions that warrant rigorous investigation [11,12]. Comparative analyses of pre- and post-T&T cohorts are needed to determine whether earlier treatment reduces HIV-related vascular inflammation or inadvertently exacerbates HTN risk due to prolonged ART exposure [13]. This study aims to investigate whether specific ART regimens and how they differentially contribute to hypertension development through metabolic or drug-specific mechanisms, and examine how structural determinants like healthcare access, socioeconomic disparities, and HIV-related stigma exacerbate HTN disparities among PLWH. This study seeks to inform optimized ART selection and integrated interventions to improve both HIV and cardiovascular outcomes in the treatment and transmission (T&T) era.

## Methods

### Study Design

This was a retrospective cohort study among PLWH aged ≥15 years enrolled in HIV care in 12 President’s Emergency Plan for AIDS Relief (PEPFAR)-supported districts in one province of Zambia.

### Study setting

The study was conducted in southern province, Zambia. The research spanned 12 districts including Chikankata, Choma, Kalomo, Kazungula, Livingstone, Mazabuka, Monze, Namwala, Pemba, Siavonga, Sinazongwe, and Zimba. The ART department records were utilized due to their systematic documentation of variables in smart care [14].

### Eligibility and recruitment criteria

The primary study abstracted data from medical health records (electronic SmartCare systems and paper-based records) for People Living with HIV (PLHIV) aged ≥15 years enrolled in care across 12 PEPFAR-supported districts in Southern Province, Zambia, and excluded participants with missing essential demographic information (age, sex, or marital status). Eligible participants were required to have documented systolic and diastolic blood pressure values. We further excluded individuals lacking these blood pressure measurements. From 9,401 files initially available for review, 8,409 medical records of people living with HIV (PLWH) aged ≥15 years were screened. A total of 6,409 records met the inclusion criteria and were included in the final analysis. The remaining 2,000 records were excluded due to incomplete demographic data (e.g., missing age, sex, or marital status), while an additional 992 records lacked essential clinical information (e.g., outcome measures, diagnoses, or treatment details).

### Study Variables

The primary outcome, hypertension (HTN) among people living with HIV, was defined as systolic blood pressure (SBP) ≥140 mmHg and/or diastolic blood pressure (DBP) ≥90 mmHg [15]. Independent variables included sociodemographic characteristics (age, sex, marital status), clinical measures (SBP, DBP, height, weight), HIV-specific indicators (viral load, CD4 count, WHO staging, ART regimen), and study design factors (cohort assignment, health facility).

### Data collection

The primary study was conducted across 45 health facilities in 12 district of southern province. Clinical data for people living with HIV (PLHIV) were abstracted from HIV medical records (paper-based or electronic/SmartCare systems) across 45 health facilities in 12 districts of Southern Province, using the Electronic Data Capture (REDCap) platform. Outcomes for PLHIV were systematically documented. In facilities where SmartCare was not fully implemented, supplementary clinical data including ART regimen, WHO stage, BMI, duration on ART, retention status, laboratory parameters such as CD4 count, viral load and pharmacy records were sourced directly from laboratory and pharmacy registers. Where feasible, data triangulation between SmartCare and paper-based sources was performed to enhance completeness and accuracy [16]. A review of medical records was conducted from November 10, 2024, to December 19, 2024.

### Data analysis

The data extracted from the REDCap platform were cleaned using Microsoft Excel before analysis in StatCrunch. Descriptive statistics included frequencies and percentages for categorical variables, while continuous variables were summarized using medians with interquartile ranges. Normality of data distribution was assessed using the Shapiro-Wilk test. For comparing median values between groups, we employed the Wilcoxon rank-sum test, while associations between categorical variables were examined using chi-square tests. To identify factors associated with HTN in BTT and ATT cohort, we performed multivariable logistic regression analysis. The selection of covariates - including age, sex, facility location, marital status, WHO clinical stage, ART regimen, and BMI - was informed by existing literature [15] and expert knowledge in HIV care and management. These variables were included in the final adjusted model based on their established relevance in prior research. Statistical significance was determined at p<0.05 for all analyses.

### Ethical considerations and consent to participants

Ethical approval for the data used in this study was obtained from the Mulungushi University School of Medicine and Health Sciences (SOHMS) Research Ethics Committee (ethics Reference number SMHS-MU2-2024-86) on 23^rd^ July 2024. No information leading to identification of patients during and after analysis was abstracted and entered in the data collection form. Secondary data were used in this project. A written/verbal consent was not applicable and was therefore waived by the ethics committee.

## Results

From 9,401 files initially available for review, 8,409 medical records of people living with HIV (PLWH) aged ≥15 years were screened. A total of 6,409 records met the inclusion criteria and were included in the final analysis. The remaining 2,000 records were excluded due to incomplete demographic data (e.g., missing age, sex, or marital status), while an additional 992 records lacked essential clinical information (e.g., outcome measures, diagnoses, or treatment details).

### Basic characteristics of study participants

The study included 6,409 participants, with 45.6% (n = 2,920) in the before Test-and-Treat (BTT) cohort and 54.4% (n = 3,489) in the after Test-and-Treat (ATT) cohort, **Table 1**. The median age of participants was higher in the BTT cohort (40 years [IQR: 34-47]) compared to the ATT cohort (37 years [IQR: 30-44]), **Figure 1**. A majority of participants in both groups were female, comprising 60.6% in BTT and 60.0% in ATT. The proportion of married individuals was slightly higher in the ATT cohort (14.9%) than in the BTT cohort (11.1%). Urban residents predominated in both groups, although the proportion was higher in the BTT cohort (93.6%) compared to the ATT cohort (89.1%). Conversely, the ATT cohort had a greater proportion of rural participants (11.0% vs. 6.4%). Systolic and diastolic blood pressures were slightly higher in the ATT group (120 mmHg and 75 mmHg, respectively) compared to the BTT group (119 mmHg and 74 mmHg, respectively).The median body mass index (BMI) was slightly lower in the ATT group (20.4 kg/m² [IQR: 10.0-23.4]) compared to the BTT group (21.0 kg/m² [IQR: 18.5–23.9]). There was a shift in ART regimens over time: the use of NNRT/NRTI-based regimens declined from 70.5% in the BTT cohort to 56.1% in the ATT cohort, while the use of integrase strand transfer inhibitors (INSTIs) increased from 26.3% to 41.5%. The use of protease inhibitors and other regimens remained low in both groups. Most participants were classified as WHO Stage I at baseline in both the BTT (94.8%) and ATT (93.3%) cohorts. Median viral load values were similar between groups, with 20 copies/mL in both cohorts. The median CD4 count was slightly higher in the BTT cohort (495 cells/µL [IQR: 327-685]) compared to the ATT cohort (470 cells/µL [IQR: 306–672]). Baseline creatinine levels were higher in the ATT cohort (73 µmol/L [IQR: 60-87.5]) than in the BTT cohort (69 µmol/L [IQR: 54.7-84.1]). Hemoglobin levels were similar between groups, with a median of 13 g/dL in both. Alanine transaminase (ALT) levels were marginally lower in the ATT cohort (24 U/L [IQR: 17–33]) than in the BTT cohort (25.6 U/L [IQR: 19– 35.1]) respectively.

**Figure 1.**
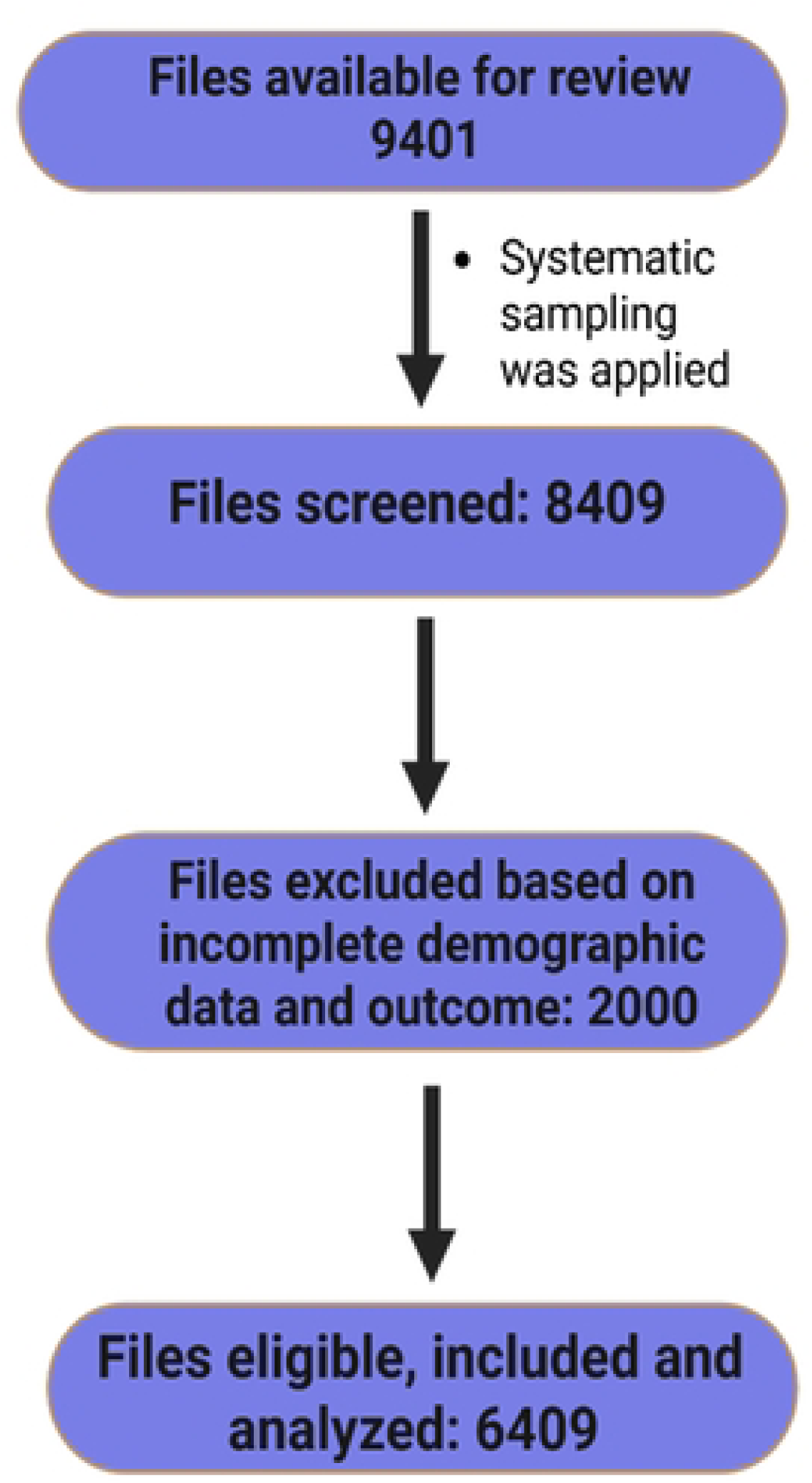
Participant recruitment flow.

**Table 1.** Characteristics of study population.

| Variable | Before test-and-treat<br>n= 2920 (45. 6%) | After test-and-treat<br>n= 3489 (54.4%) | P value |
| --- | --- | --- | --- |
| **Sex** |  |  |  |
| Male | 1151 (39.4) | 1397 (40.0) | 0.602 |
| Female | 1769 (60.6) | 2092 (60.0) |  |
| **Marital status** |  |  |  |
| Married | 323 (11.1) | 560 (14.9) | **<0.0001** |
| Unmarried | 2597 (88.9) | 2969 (85.1) |  |
| **Facility** |  |  |  |
| Rural | 52 (6.4) | 44 (11.0) | **0.0001** |
| Urban | 762 (93.6) | 358 (89.1) |  |
| **ART Regimen** |  |  |  |
| NNRT/NRTIs | 2061 (70.5) | 1956 (56.1) | **<0.0001** |
| INSTIs | 768 (26.3) | 1149 (41.5) |  |
| PIs | 84 (2.9) | 77 (2.2) |  |
| Other | 7 (0.2) | 7 (0.2) |  |
| **WHO clinical staging of HIV** |  |  |  |
| 1 | 2771 (94.8) | 3258 (93.3) | **< 0.001** |
| 2 | 29 (0.9) | 22 (0.6) |  |
| 3 | 18 (0.6) | 12 (0.3) |  |
| 4 | 3 (3.3) | 2 (0.05) |  |
| **Abbreviation:** IQR; interquartile range, NNRTI; non-nucleoside/nucleotide reverse transcriptase inhibitor, PI; protease inhibitor, INSTI; integrase strand transfer inhibitor, NRTI; nucleotide reverse transcriptase inhibitor. |  |  |  |

### Relationship between demographic, clinical characteristic and Hypertension

The cumulative incidence of hypertension was 8.8% (258/2920) before the test-and-treat policy and increased to 10.2% (355/3489) after its implementation, **Table 2**. A higher proportion of individuals with hypertension were observed in urban areas compared to rural areas (43.0% vs. 57.0% after test-and-treat; 29.2% vs. 70.8% before test-and-treat). Individuals with hypertension were generally older compared to those without hypertension, with median ages of 46 vs. 40 years before the test-and-treat policy and 44 vs. 36 years after, p<0.0001, **Figure 2**. A higher proportion of males had hypertension compared to females in both periods, 10.6% vs. 7.7% before test-and-treat and 12.2% vs. 8.8% after, p=0.0067 and p=0.0015, respectively. A higher proportion of individuals who initiated ART before the implementation of the test-and-treat policy had hypertension (82.1% vs. 17.9%). Additionally, individuals with hypertension had a higher baseline hemoglobin level, body mass index, and creatinine levels compared to those without hypertension. Other clinical markers such as viral load and CD4 count showed no significant differences between hypertensive and non-hypertensive individuals respectively.

**Figure 2.**
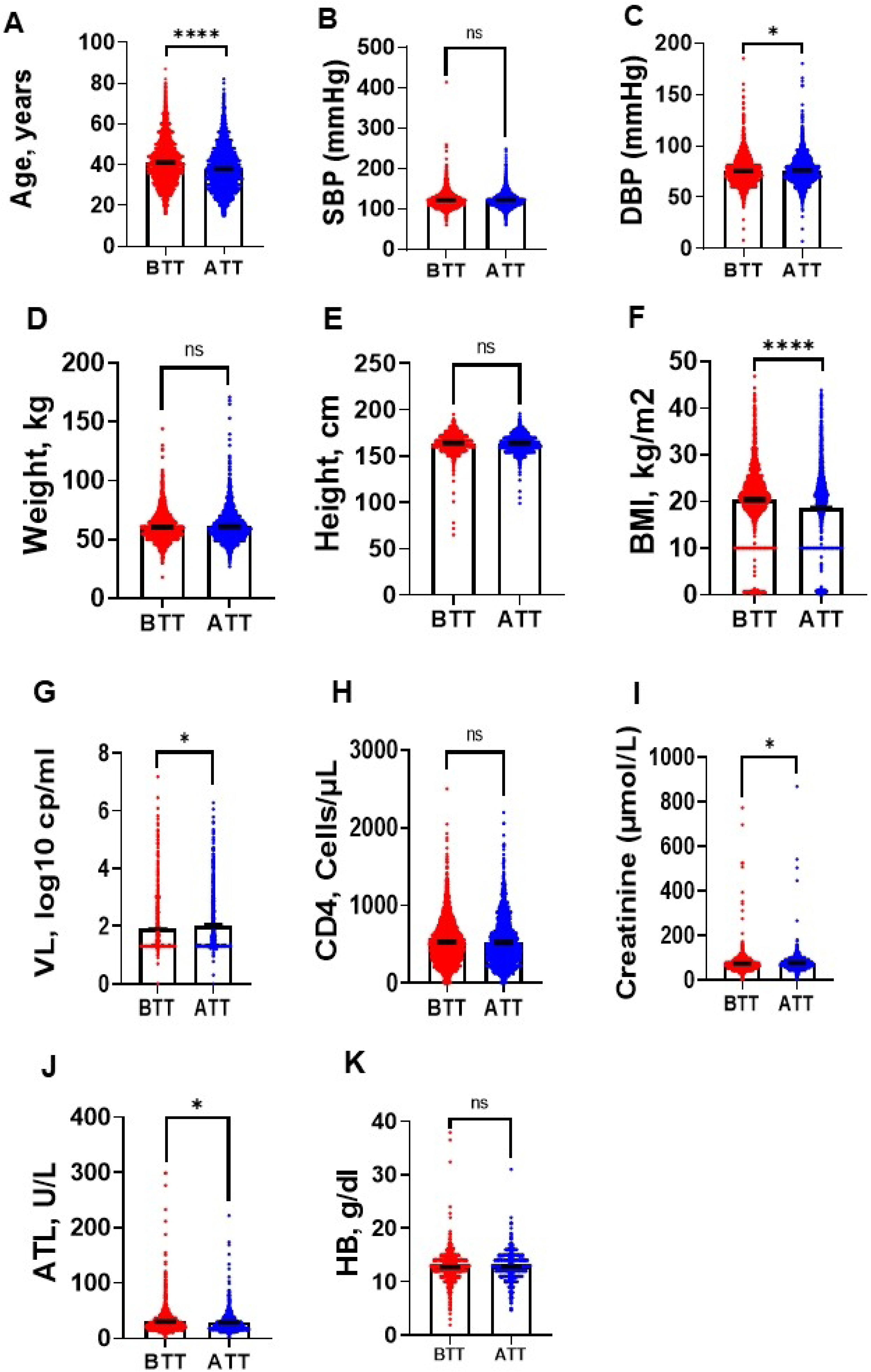
Blood and anthropometric parameters between the before test-and-treat cohort and the after test-and-treat cohort. A. Age between before test and treat (n= 2920) and after test and treat cohort (n= 3489). B. systolic blood pressure between before test (n= 2737) and treat and after test and treat cohort (n= 3216). C. diastolic blood pressure between before test and treat (n= 2738) and after test and treat cohort (n= 3217). D. Weight between before test and treat (n= 2653) and after test and treat cohort (n= 2815). E. height between before test and treat (n= 2528) and after test and treat cohort (n= 2569). F. BMI between before test and treat (n= 2916) and after test and treat cohort (n= 3815). G. Viral load between before test and treat (n= 806) and after test and treat cohort (n= 953). H. CD4 count between before test and treat (n= 2245) and after test and treat cohort (n= 1476). I. Creatinine between before test and treat (n= 1615) and after test and treat cohort (n= 973). J. ALT between before test and treat (n= 1540) and after test and treat cohort (n= 912). K. HB between before test and treat (n= 1765) and after test and treat cohort (n= 1071).*p<0.05; **p<0.01; ****p<0.001ns, p>0.05.

**Figure 3.**
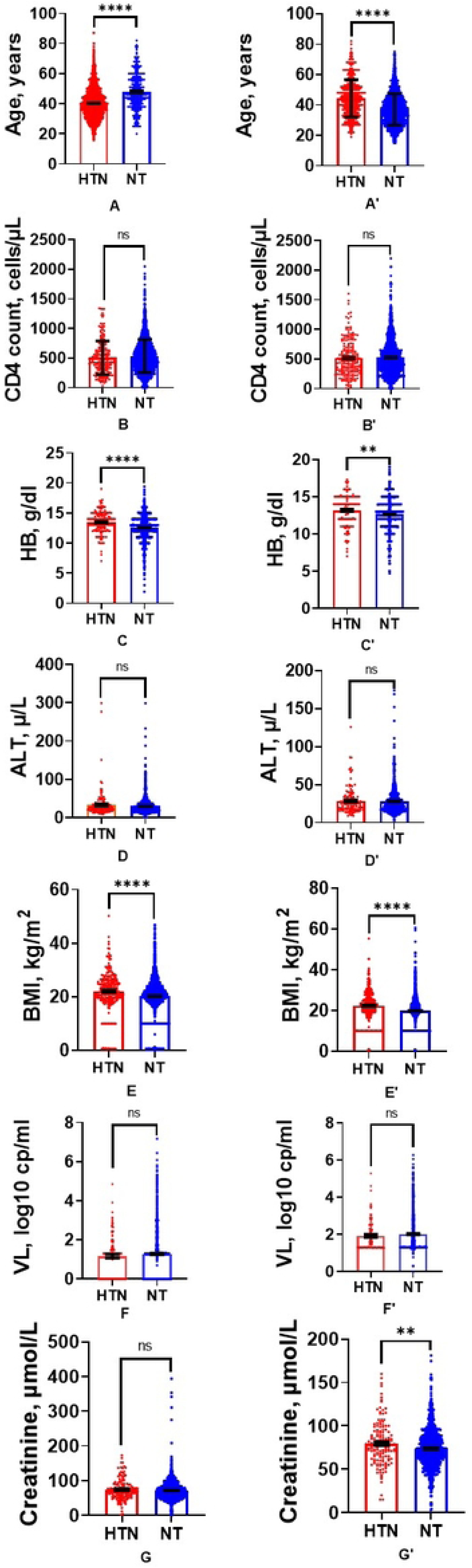
Laboratory and clinical parameters of Hypertensive and normotensives for both before and after test and treat policy. A. Age of hypertensive (n= 258) and normotensives (n= 2662) in before test and A’) hypertensive (n= 355) and normotensives (n= 3134) in the after test and treat cohort. B. CD4 count of hypertensive (n= 203) and normotensives (n= 2042) in before test and B’) hypertensive (n= 196) and normotensives (n= 1280) after test and treat. C. HB of hypertensive (n= 151) and normotensives (n= 920) in before test and treat cohort and C’) hypertensive (n= 157) and normotensives (n= 1608) in the after test and treat cohort. D. ALT of hypertensive (n= 131) and normotensives (n= 781) in before test and treat and D’) hypertensive (n= 144) and normotensives (n= 1396) after test and treat. E. BMI of hypertensive (n= 257) and normotensives (n= 2659) in before test cohort and E’) hypertensive (n= 335) and normotensives (n= 2878) after test and treat. F. Viral Load of hypertensive (n= 107) and normotensives (n= 846) in before test and treat and F’) hypertensive (n= 112) and normotensives (n= 1100) after test and treat. G. Plasma creatinine of hypertensive (n= 156) and normotensives (n= 1459) in before test and treat cohort and G’) hypertensive (n= 147) and normotensives (n= 826) after test and treat. (n= 1071). *p<0.05; **p<0.01; ****p<0.001; ns, p>0.05.

**Table 2:** Relationship between demographic, clinical characteristic and Hypertension.

| Variable | Before test-and-treat |  |  |  | After test-and-treat |  |  |  |
| --- | --- | --- | --- | --- | --- | --- | --- | --- |
|  | Median (IQR) or Frequency (%) | HTN | NT |  | Median (IQR) or Frequency (%) | HTN | NT |  |
|  |  | Yes=258 (8.8%) | No =2662 (91.2%) | P-value |  | Yes=355 (10.2%) | No =3134 (89.8%) | P-value |
| **Sex** |  |  |  | **0.0067** |  |  |  | **0.0015** |
| *Male* | 1151 (39.4) | 122 (10.6) | 1029 (89.4) |  | 1397 (40.0) | 170 (12.2) | 1227 (87.8) |  |
| *Female* | 1769 (60.6) | 136 (7.7) | 1633 (92.3) |  | 2092 (60.0) | 185 (8.8) | 1907 (91.2) |  |
| **Marital status** |  |  |  | **0.0025** |  |  |  | 0.887 |
| *Married* | 323 (11.1) | 14 (4.3) | 309 (95.7) |  | 520 (14.9) | 34 (6.5) | 486 (93.5) |  |
| *Unmarried* | 2,597 (88.9) | 244 (9.4) | 2353 (90.6) |  | 2,969 (85.1) | 321 (10.8) | 2648 (89.2) |  |
| **Residence** |  |  |  | **0.0394** |  |  |  | **0.0027** |
| *Rural* | 2,067 (70.8) | 179 (9.5) | 1870 (90.5) |  | 1,990 (57.0) | 229 (11.5) | 1761 (88.5) |  |
| *Urban* | 853 (29.2) | 61 (7.2) | 792 (92.8) |  | 1,499 (43.0) | 126 (8.4) | 1373 (91.6) |  |
| **ART** |  |  |  | 0.323 |  |  |  | 0.127 |
| *NNRT/NRTIs* | 2,061<br>(70.6) | 171 (8.3) | 1890<br>(91.7) |  | 1,956<br>(56.1) | 194<br>(10.0) | 1762<br>(90.0) |  |
| *INSTIs* | 768 (26.3) | 79 (10.3) | 689<br>(89.7) |  | 1,449<br>(41.5) | 146<br>(10.0) | 1303<br>(90.0) |  |
| *PIs* | 84 (2.9) | 8 (9.5) | 76 (90.5) |  | 77 (2.2) | 14<br>(18.2) | 63<br>(81.8) |  |
| *Other* | 7 (2) | 0 (0.0) | 7 (100.0) |  | 7 (0.2) | 1 (14.3) | 6 (85.7) |  |
| **WHO clinical staging of HIV** |  |  |  | 0.1544 |  |  |  | 0.565 |
| 1 | 2,771<br>(94.9) | 248 (9.0) | 2523<br>(91.0) |  | 3,258<br>(93.4) | 2917<br>(89.5) | 341<br>(10.5) |  |
| 2 | 29 (1) | 4 (13.8) | 25 (86.2) |  | 22 (0.6) | 20<br>(90.9) | 2 (9.1) |  |
| 3 | 18 (0.6) | 0 (0.0) | 18<br>(100.0) |  | 12 (0.2) | 11<br>(91.7) | 1 (8.3) |  |
| 4 | 3 (0.1) | 1 (33.3) | 2 (66.7) |  | 2 (0.1) | 2<br>(100.0) | 0 (0.0) |  |
| **Abbreviation.** IQR; interquartile range, NNRTI; non-nucleoside/nucleotide reverse transcriptase inhibitor, PI; protease inhibitor, INSTI; integrase strand transfer inhibitor, NRTI; nucleotide reverse transcriptase inhibitor. |  |  |  |  |  |  |  |  |

### Logistic regression of factors associated with Hypertension

Table 3 shows the results of univariable and multivariable regression analyses of factors associated with hypertension. At univariable analysis, participants who were older had 1.06 times higher odds of having hypertension than younger participants (Odds ratio (OR): 1.06, 95% CI: 1.05-1.07, p=0.0001). Participants from urban facilities had 27% lower odds of having hypertension compared to those from rural facilities (OR: 0.73, 95% CI: 0.54-0.98, p=0.040). At multivariable analysis, for every year increase in age, participants had 6% higher odds of having hypertension (adjusted odds ratio (AOR): 1.06, 95% CI: 1.04-1.07, p<0.0001). Urban residence remained significantly protective, with urban participants having 28% lower odds of hypertension compared to rural participants (AOR: 0.72, 95% CI: 0.54-0.98, p=0.041). After implementation of the test-and-treat intervention, age remained significantly associated with hypertension in both univariable (OR: 1.05, 95% CI: 1.05-1.07, p=0.0001) and multivariable analyses (AOR: 1.05, 95% CI: 1.04-1.07, p<0.0001). The protective effect of urban residence strengthened post-intervention, with urban participants having 33% lower odds of hypertension (AOR: 0.67, 95% CI: 0.53-0.85, p=0.001) compared to rural participants. Hemoglobin levels became marginally significant in the post-intervention multivariable analysis (AOR: 1.08, 95% CI: 1.00-1.12, p=0.049)

**Table 3:** Logistic regression analysis of current demographic and clinical characteristics associated with hypertension.

| Variable | Before test-and-treat |  |  |  | After test-and-treat |  |  |  |
| --- | --- | --- | --- | --- | --- | --- | --- | --- |
|  | Univariable analysis |  | Multivariable analysis |  | Univariable analysis |  | Multivariable analysis |  |
|  | OR (95 %CI) | P-value | OR (95 %CI) | P-value | OR (95%CI) | P-value | OR (95 %CI) | P-value |
| **Age** | 1.06 (1.05, 1.07) | **0.0001** | 1.06 (1.04, 1.07) | **<0.0001** | 1.05(1.05,1.07) | **0.0001** | 1.05 (1.04, 1.07) | **<0.0001** |
| **Sex** |  |  |  |  |  |  |  |  |
| *Male* | REF |  | REF |  | REF |  | REF |  |
| *Females* | 0.70 (0.54, 0.90) | **0.007** | 0.85 (0.65, 1.10) | 0.053 | 0.70(0.56,0.87) | 0.0015 | 0.84 (0.66, 1.05) | 0.133 |
| **HB** | 1.00 (0.99, 1.00) | 0.070 | 1.00 (0.99, 1.01) | 0.606 | 1.08 (1.01,1.16) | 0.202 | 1.08 (1.00, 1.12) | **0.049** |
| **BMI** | 1.00 90.99, 1.00) | 0.529 | 1.001(0.99,1.004) | 0.277 | 1.0 (0.99, 1.00) | 0.296 | 1.001(0.99,1.003) | 0.325 |
| **Marital status** |  |  |  |  |  |  |  |  |
| *Unmarried* | REF |  |  |  | REF |  |  |  |
| *Married* | 2.28 (1.31, 3.97) | 0.0033 | 1.20 (0.67, 2.14) | 0.524 | 1.73 (1.20,2.49) | 0.0033 | 0.98 (0.66, 1.44) | 0.930 |
| **Facility** |  |  |  |  |  |  |  |  |
| *Rural* | REF |  | REF |  | REF |  | REF |  |
| *urban* | 0.73 (0.54, 0.98) | 0.040 | 0.72(0.543, 0.98) | **0.0413** | 0.7- (0.56, 0.88) | 0.0028 | 0.67(0.53,0.85) | **0.0013** |
| **Abbreviations:** OR; odds ratio, AOR; Adjusted odds ratio, HB; haemoglobin; BMI, body mass index, kg/m<sup>2</sup> and REF; reference |  |  |  |  |  |  |  |  |

## Discussion

Our study aimed at finding the factors associated with HTN in PLHIV before test and treat and after test and treat. The study found a significant association between increasing age and hypertension risk among PLHIV, with each additional year of age increasing hypertension odds by 5-6% in both pre- and post-intervention periods. This finding aligns with multiple epidemiological studies demonstrating age as an independent risk factor for hypertension, likely due to age-related vascular stiffness, endothelial dysfunction, and reduced renal function [17,18].The consistent association across both study periods suggests that aging processes may outweigh the potential protective effects of ART initiation under the test-and-treat policy [19]. This suggests the need for intensified blood pressure monitoring and cardiovascular risk assessment in older PLHIV, particularly as improved ART has increased life expectancy in this population [20,21].

Another interesting finding was hemoglobin levels. Hemoglobin levels showed an association with hypertension risk post-intervention, with each unit increase in hemoglobin marginally increasing hypertension odds. This finding may reflect several potential mechanisms. Higher hemoglobin could indicate improved nutritional status following antiretroviral therapy (ART) initiation, potentially associated with weight gain, increased red cell mass, and metabolic alterations [22,23]. Alternatively, it may suggest increased blood viscosity contributing to elevated peripheral vascular resistance and subsequent blood pressure rise [24]. The absence of this association prior to ART introduction suggests the relationship may be mediated by ART-induced physiological changes [25]. Furthermore, recent studies have begun to explore these links more closely. A 2024 study by Gudina et al. reported that hemoglobin elevation following ART initiation was significantly correlated with increases in systolic blood pressure, particularly among patients on integrase and Dolutegravir based regimens [26] Similarly, Denu et al. (2024) found that ART-associated hematologic recovery was linked to an increased risk of developing metabolic syndrome, including hypertension, among women living with HIV [27]. Additionally, Nguyen et al. noted that ART-induced erythropoiesis and shifts in inflammatory cytokine profiles could influence vascular tone and contribute to blood pressure dysregulation [28]. These findings suggest that although ART effectively restores immune and hematological function, it may also alter cardiovascular risk profiles in ways that require deeper exploration especially regarding hematologic effects specific to different treatment regimens.

Another variable worth discussing was residence. Urban residence demonstrated a protective effect against hypertension, with urban participants having 28-33% lower odds compared to rural residents. This finding contrasts with conventional urban health paradigms that typically associate urban environments with increased cardiovascular risk factors[29].The observed protection may reflect better healthcare access in urban centers, including earlier HIV diagnosis, more consistent ART availability, and greater hypertension screening capacity [30]. However, this protective association warrants cautious interpretation, as urban-rural differences in healthcare-seeking behavior, dietary patterns, or physical activity levels were not assessed in this study. The strengthening of this protective effect post-intervention from 28% to 33% lower odds may indicate that urban health systems were better positioned to implement the test-and-treat policy effectively.

### 4.2 Strengths and limitations

The study demonstrates notable strengths, including a large, representative sample of 6,409 participants across 12 Zambian districts, enhancing statistical power and generalizability to PLHIV in similar sub-Saharan African contexts, as well as a robust pre- and post-T&T cohort comparison (BTT and ATT) that provides critical insights into hypertension risk shifts following updated HIV treatment guidelines. Comprehensive data integration from electronic (SmartCare) and paper-based records minimized information bias, while systematic capture of clinical variables (e.g., ART regimens, blood pressure, CD4 counts) supported rigorous analysis, including the novel finding of urban residence being protective against hypertension, suggesting healthcare access advantages. Additionally, its focus on the transition from NNRTI/NRTI to INSTI-based regimens aligns with global ART trends, offering timely evidence on cardiovascular risks linked to modern therapies. However, limitations include the retrospective design’s susceptibility to missing data and documentation errors, exemplified by the exclusion of 2,000 incomplete records, which may introduce bias. The cross-sectional approach and lack of longitudinal follow-up preclude causal conclusions between ART and hypertension, while unmeasured confounders (diet, physical activity, genetic factors) risk residual confounding. Although viral loads were similar between cohorts, minor statistical differences (p=0.0024) lack clarity without adherence or resistance data, potentially obscuring treatment-efficacy nuances relevant to hypertension risk. These limitations underscore the need for prospective studies integrating behavioral and genetic data to validate findings.

### Conclusion

This study provides valuable insights into the shifting burden of hypertension (HTN) among people living with HIV (PLHIV) in Zambia, particularly before and after the implementation of the universal test-and-treat (T&T) policy. The findings indicate a notable rise in HTN prevalence following the T&T policy, increasing from 8.8% to 10.2%. This trend highlights the growing cardiovascular risks within this population as access to antiretroviral therapy (ART) expands. Age consistently emerged as a significant predictor of HTN, with each additional year increasing the likelihood by 5– 6%. This underscores the heightened vulnerability of older PLHIV, likely due to age-related vascular changes and prolonged exposure to ART. Interestingly, urban residence appeared to have a protective effect against HTN, possibly due to better healthcare access, earlier ART initiation, and more consistent hypertension screening. These finding challenges conventional assumptions that urban environments contribute to higher cardiovascular risk and suggests that improved health infrastructure can help mitigate HTN among PLHIV. Additionally, the observed marginal association between elevated hemoglobin levels and HTN post-T&T raises important questions about ART-induced metabolic and hematologic changes that may influence blood pressure regulation. Despite the study’s strengths, including a large and representative sample with robust data collection, its retrospective design and lack of longitudinal follow-up limit causal interpretations. Unmeasured factors such as lifestyle habits (diet, physical activity and smoking) and genetic predisposition may also play a role in the observed associations. Nonetheless, these findings highlight the urgent need for integrated HTN screening and management within HIV care programs, particularly for aging populations and rural communities with limited healthcare access. The transition to INSTI-based regimens post-T&T, although not directly linked to HTN in this study, warrants further investigation into their long-term metabolic effects. Future research should adopt prospective designs to better understand the causal relationships between ART, immune recovery, and cardiovascular risk. Policymakers and healthcare providers must prioritize routine blood pressure monitoring, lifestyle interventions, and targeted HTN management strategies to address the dual burden of HIV and cardiovascular disease in sub-Saharan Africa. In conclusion, while the T&T policy has significantly improved HIV outcomes, complementary strategies are essential to tackle the rising HTN epidemic among PLHIV. Ensuring that longer survival is accompanied by enhanced cardiovascular health should be a key priority in HIV care programs.

## Availability of data and materials

The raw data underlying the results presented in the study have been uploaded as supporting information.

## Author’s contributions

SKM, AK and MC conceived the Study. SKM, AK and MC oversaw Data acquisition. SKM, AK, MC, and SL supervised data acquisition. MC and DC conducted the formal analysis. MC, DC, SL, JPP, BMH, AK and SKM wrote the original draft. All authors contributed to the article edits and approved the final manuscript.

## Acknowledgements

We want to thank the HAND research group members from Mulungushi University School of Medicine and Health Sciences for their support in data collection. We used graph pad prism to draw the figure in the discussion.

S1. Strobe checklist

S2. Dataset

